# Federated Multiple Imputation for Variables that Are Missing Not At Random in Distributed Electronic Health Records

**DOI:** 10.1101/2024.09.15.24313479

**Authors:** Yi Lian, Xiaoqian Jiang, Qi Long

## Abstract

Large electronic health records (EHR) have been widely implemented and are available for research activities. The magnitude of such databases often requires storage and computing infrastructure that are distributed at different sites. Restrictions on data-sharing due to privacy concerns have been another driving force behind the development of a large class of distributed and/or federated machine learning methods. While missing data problem is also present in distributed EHRs, albeit potentially more complex, distributed multiple imputation (MI) methods have not received as much attention. An important advantage of distributed MI, as well as distributed analysis, is that it allows researchers to borrow information across data sites, mitigating potential fairness issues for minority groups that do not have enough volume at certain sites. In this paper, we propose a communication-efficient and privacy-preserving distributed MI algorithms for variables that are missing not at random.

## Introduction

Electronic health records (EHR) have been widely implemented and utilized in healthcare. Nationwide EHRs for England, Wales, Scotland, Denmark, and Sweden have been used in research for years^1^. The linked EHR research environment for England contains EHRs from primary care, hospital episodes, death registry and others for more than 50 million people, accounting for over 96% of the English population^1^. With access to such population-wide resources, researchers are seeing great opportunities, as well as significant challenges. Given the massive sizes of these EHRs, it may be more reasonable, feasible and efficient to store data locally or at multiple data centers instead of a central repository. Similarly, being able to take advantage of distributed computing resources may become necessary in order to perform the desired analyses. More importantly, in the presence of restrictions and policies regarding data sharing, generally centered around privacy concerns, data from different sources may not be pooled together. For example, the Patient-centered Scalable National Network for Effectiveness Research (pSCANNER) features a distributed architecture containing data from 13 sites covering over 37 million patients^2^. Privacy-preserving distributed learning or federated learning methods have been developed and proven to be effective in practice^3–7^. We use the term “distributed” in the remainder of the paper. Dedicated distributed analyses can also contribute to fairness in machine learning. In contrast to the vast total size of the distributed data, some sites can have limited amount of samples, particularly for certain minority groups or rare diseases. Without adequate volumes of data, the accuracy and reliability of inference and prediction results can be compromised. This issue can be alleviated by borrowing strength from the same minority groups from other sites, through distributed data analysis.

Missing data is common in EHRs, and naturally, distributed EHRs as well. Researchers can use various imputation methods to handle missing data^8^ but missing data problems in distributed EHRs can be more challenging. Particularly, the proportion, pattern and even mechanism of missingness of the same variable may vary greatly from site to site, as a result of local regulations, legislation and even culture and demographics. We now use the Georgia Coverdell Acute Stroke Registry (GCASR) data we analyze later in this paper as an example. EHR collected from a number of participating hospitals are treated as distributed EHR in our analysis for demonstration purpose. We aim to estimate the association between the arrival-to-computed tomography (CT) time (an important indicator of acute stroke care quality^9^) and whether the hospital receives advance notification from the emergency medical services (EMS) before the stroke patient arrives^10^ (EMSNote) adjusting for potential confounders including Gender and Weekend (arrived at the hospital on weekends instead of weekdays). The binary EMSNote is subject to missingness and the imputation of such variables under distributed setting can be more difficult to address than in centralized data in a few ways. We plot the distribution of missing proportions of EMSNote at all hospitals in Figure 1. While most sites have under 2% missing that may not be a significant issue, the proportion can be quite high in a number of sites that are generally considered challenging to impute locally (e.g. Hospital B in Figure 1 with 43% missing and Hospital C with 89%). In addition, some sites may have very small sample sizes (e.g. Hospital A with 9 records) such that reliable imputation cannot be performed locally either. In summary, under distributed settings, sites can have small data volumes, high proportions of missingness or both that are harmful to imputation performance. These issues do not exist in the much larger aggregated GCASR dataset (overall 7.8% missing) and are the motivations for distributed imputation algorithms when pooling is not feasible. In addition, the probability of missingness could be higher in some groups than others, thus disproportionally compromise the imputation and subsequent analysis performance in some groups, causing fairness issues. Therefore, communication-efficient and privacy-preserving distributed MI methods (that can facilitate borrowing information across sites) are needed to safeguard the distributed analyses of distributed EHRs.

**Figure 1.**
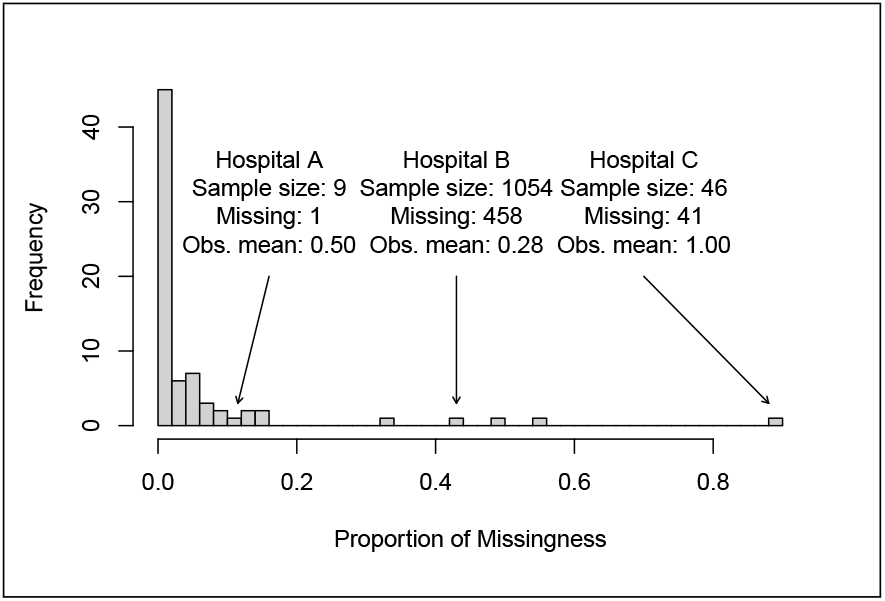
Proportions of missingness of EMSNote (whether advance notification of stroke by emergency medical services is received) collected at participating hospitals in the Georgia Coverdell Acute Stroke Registry. Highlighted are examples of challenging characteristics for local imputation such as small sample size (Hospital A), high missing proportions (Hospitals B & C) and evidence of potential missing not at random (all observed values equal to 1 at Hospital C).

To our knowledge, there is limited literature on distributed MI methods. The most relevant work focuses on missing at random (MAR) mechanism^11^, one of the three widely-recognized missing data mechanisms along with missing completely at random (MCAR) and missing not at random (MNAR). MCAR refers to the cases where the probability of missing is the same for all observations and is unrelated to the data^8^. In the example in Figure 1, the low missing proportions observed in most hospitals might be attributed to random human errors thus could be considered MCAR. However, one would reasonably suspect that some systematic reasons are causing, for example, Hospital B in Figure 1 to not record EMSNote 43% of the time. In the MAR cases, the probability of being missing may depend on and the missingness can be accounted for by observed data^8^. In the MNAR case, the probability is effected by unobserved factors, such as the latent value of the missing variable itself or unknown correlation between the latent value of the missing variable and the missing mechanism^8^. For instance, Hospital C in Figure 1 is missing 89% of its EMSNote information and the observed values are uniformly equal to 1. This suggests that, potentially, this hospital only records EMSNote when the value is 1 thus the missingness is strongly associated with EMSNote being equal to 0. In practice, it is generally difficult, if possible at all, to test whether an incomplete variable is MAR or MNAR^8^. Therefore, there is need for robust distributed MI methods that can effectively impute MAR and MNAR data, which is the main contribution of this work. Based on existing Heckman imputation models for non-distributed data^12^ and a distributed MI framework^11^, we present HDMI (Heckman distributed multiple imputation) – a communication-efficient and privacy-preserving MI method for distributed EHRs. The HDMI works by fitting imputations models in a distributed manner such that the sites that cannot generate reliable and accurate local imputation results can borrow information from other sites without sharing individual patient data.

## Methods

We continue to use the GCASR data as an example to help explain the method. Assume that we use a linear model to estimate the association between a continuous outcome variable *Y* ∈ *ℝ*^*N*^ and a set of covariates **X** = (1, *X*_1_, …, *X*_*p*_) ∈ ℝ^*N×*(*p*+1)^. Data is distributed at *k* = 1, …, *K* different sites (hospitals), and the site-specific datasets are denoted by *Y* ^(*k*)^ and **X**^(*k*)^ with sample size *n*^(*k*)^ such that ∑_*k*_ *n*^(*k*)^ = *N*. We refer to the model

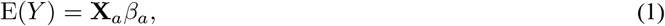

as the “analysis model”, where 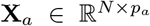 is a subset of **X** involved in the analysis model and 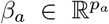 is a vector of coefficients (subscript *a* for “analysis”). The goal of the analysis is to estimate *β*_*a*_, which quantifies the association between the outcome variable *Y* and covariates of interest. In our example, we estimate the coefficients in E(Arrival to CT Time) = *β*_*a*0_ + *β*_*a*1_EMSNote + *β*_*a*2_Weekend + *β*_*a*3_Gender. Assume that only one variable *X*_1_ (EMSNote in our example) has missing values denoted by *X*_1,mis_ along with observed values *X*_1,obs_. Let *R* denote the missing indicator of *X*_1_, where *R*_*i*_ = 1 if *X*_1*i*_ ∈ *X*_1,obs_ and *R*_*i*_ = 0 if *X*_1*i*_ ∈ *X*_1,mis_ for the *i*-th observation, *i* = 1, …, *n*. If *X*_1_ is MAR or MNAR, proper imputation procedure is needed, otherwise we will acquire biased estimates of *β*_*a*_. Following a previous work on distributed MI and analysis^11^, our work consists of distributed imputation, distributed analysis and pooling, similar to ordinary non-distributed MI methods^8^.

### Distributed imputation

A Heckman sample selection model^13,14^ is used as the basis for the imputation model following previous work^12,15^. Below we provide a gentle introduction of the Heckman model, which addresses MNAR data problems by jointly estimating a “selection model” and an “outcome model” (the missing variable is the “outcome”). The selection model below aims to associate the binary missingness status with the observed variables in each distributed dataset,

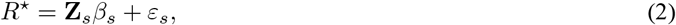

where *R*^⋆^ is a latent continuous variable that determines the binary missing indicator *R* through *R* = 1 if *R*^⋆^ *>* 0 and *R* = 0 otherwise, 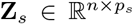 denotes the subset of variables in (*Y*, **X**), not including *X*_1_ itself, that is included in the selection model and 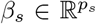 are the selection model coefficients (subscript *s* for “selection”). The outcome model quantifies the association between the missing variable and observed variables based on observed data. In our case,

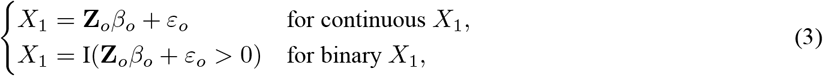

where 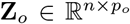 is the subset of variables in (*Y*, **X**) used, 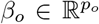 are the outcome model coefficients and I(·) is the indicator function (subscript *o* for “outcome”). It is recommended that the selection model includes at least one extra variable to avoid collinearity issues, known as the exclusion-restriction rule^16^. In our example, the selection and outcome models are fitted using an intercept, arrival-to-CT time, Weekend, and Gender, plus EducEMS (record of education on activating EMS) as the extra variable in the selection model. Heckman model assumes that the error terms of the models, *ε*_*s*_ and *ε*_*o*_, are correlated through a bivariate normal distribution, such that the value of the missing variable itself is associated with its missingness, i.e. MNAR. The Heckman model involves a correlation coefficient *ρ* as a model parameter to adapt to both missing mechanisms. When *ρ* ≠ 0, the mechanism is MNAR and larger *ρ* means stronger MNAR mechanism^12^. When *ρ* = 0 the mechanism is MAR because the value of *X*_1_ itself is neither directly associated with its missingness in (2), nor is it indirectly associated through the correlated noise terms when *ρ* = 0 is independent of *X*_1_^17^. Computation-wise, joint bivariate models can be used to estimate the selection and outcome model coefficients^18^, i.e. *θ* = (*β*_*s*_, *β*_*o*_, *ρ*) if the missing variable is binary and *θ* = (*β*_*s*_, *β*_*o*_, *ρ, σ*_*o*_) if the missing variable is continuous (the standard deviation *σ*_*o*_ of *ε*_*o*_ is assumed to be 1 for the binary case). We adopt the maximum likelihood approaches to estimate *θ* for both continuous missing variable^19^ and binary missing variable^15,18,20^, Using the estimated Heckman model parameters 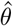 and their variance-covariance matrix 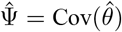, an imputation algorithm for MNAR data has been developed previously^12^ (summarized in Algorithm 1). Like other MI methods, the algorithm works by randomly drawing numbers from the probabilistic distribution of the missing variable estimated by the imputation model to fill in the missing observations. In summary, Heckman imputation model is designed for MNAR variables but also works for MAR as a special case. It is particularly useful under distributed settings when some of the sites show evidence of MNAR while others could be MAR, as in our example.

Based on Algorithm 1 and a distributed MI framework for MAR data^11^, we develop the HDMI algorithm that performs distributed MI for MNAR data. Specifically, HDMI adopts the average mixture (AVGM) approach^3^, where an imputation model is fitted at each site (hospital) *k* using only local data (*Y* ^(*k*)^, **X**_(*k*)_) and the imputation model estimates are averaged to find the global estimate. Now denote the imputation model estimates from site *k* by 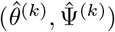. The global estimate is computed by taking an average of the site-specific estimates weighted by the number of complete cases 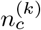 at each site^11^,

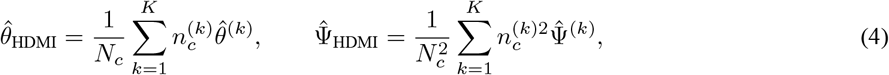

where 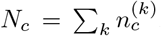. Then at each site *k*, imputation is performed by calling Algorithm 1 using the global estimate acquired in (4) to impute 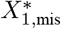. A unique seed may be used to make sure that *θ*^*^ drawn in the first step of Algorithm 1 is the same at all sites. In practice, the imputation component of the HDMI is communication-efficient as it only requires a one-way communication from the sites to the central server to deliver 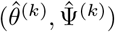 and another one-way communication from the central server to the sites for the weighted average 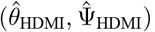. We note that 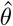 ’s are vectors of length *p*_*s*_+*p*_*o*_+2 for continuous and *p*_*s*_ + *p*_*o*_ + 1 for binary missing missing variable and 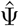’s are square matrices of the same dimension. In addition, as the communications only transmit imputation model estimates, data privacy is preserved.

#### Algorithm 1: Heckman imputation for variables that are missing not at random in a single dataset^12^

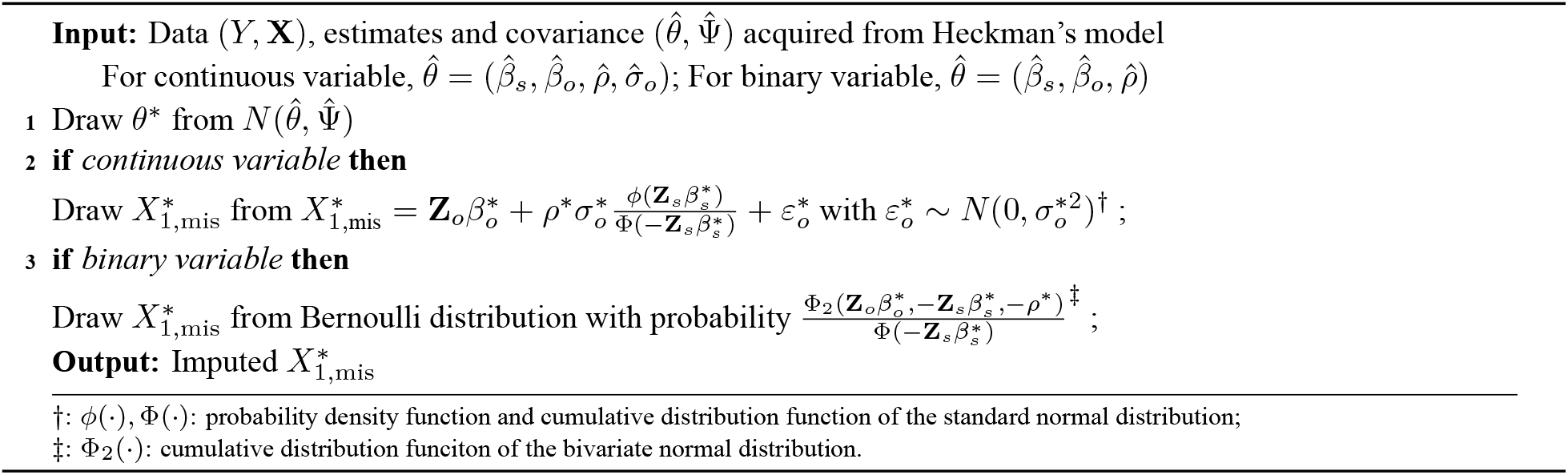

### Distributed analysis

The next component of the HDMI algorithm involves standard distributed analyses that are also communication-efficient and privacy-preserving. At each MI iteration *m*, each site (hospital) *k* is now able to generate imputed 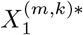 that consists of 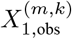 and 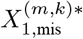 and therefore imputed matrix of analysis model predictors denoted by 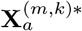. For the continuous outcome *Y*, we use distributed linear regression that only requires summary statistics from the sites^11^. The global estimates of the analysis model can be computed in closed form

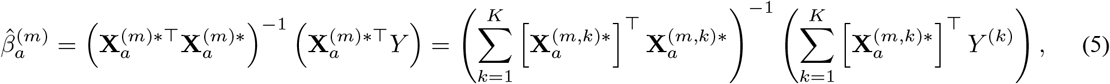

where 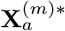 is **X**_*a*_ with imputed values, from the *m*-th imputation, that we do not have access to in the distributed learning setting. The distributed linear regression in (5) is also communication-efficient and, to some extent, privacy-preserving, due to the fact that only site-specific summary statistics (**X**^⊤^**X** and **X**^⊤^*Y*) are transmitted once to the central server. Different distributed analysis models can be used here. We use linear regression for demonstration purposes as the focus of this work is on the distributed imputation of MNAR variables.

### Pooling

In the last step, after we perform the aforementioned distributed imputation and distributed analysis procedures for a total of *M* imputation iterations, we combine the results from the analysis model to acquire the final estimates. Following Rubin’s rule^21^, the final analysis model estimates is computed by 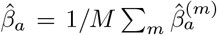. We summarize the HDMI in Algorithm 2.

#### Algorithm 2: Heckman-type Distributed Multiple Imputation algorithm (HDMI).

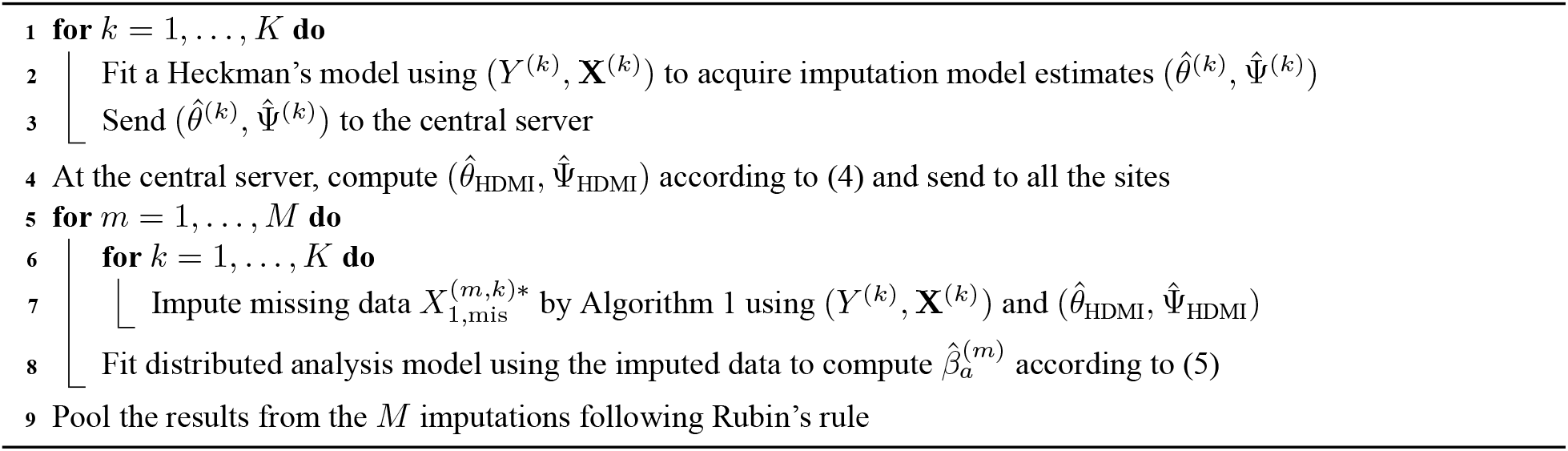

## Results

### Simulation

We perform a series of simulation analyses to test the performance of the HDMI algorithm. We consider three settings with different missing mechanisms, namely a) MAR, b) Heckman-type MNAR and c) non-Heckman MNAR^12^. There are a total of four variables *Y, X*_1_, *X*_2_ and *X*_3_, where *X*_1_ and *X*_2_ are predictors of *Y* in the analysis model, and *X*_3_ is the additional variable associated with the missingness of *X*_1_ in the true selection model (as per the exclusion-restriction rule). Both *X*_2_ and *X*_3_ are randomly generated from a normal distribution *N* (0, 0.5^2^). In all three settings, we consider the following true analysis model *Y* = 1 + *X*_1_ + *X*_2_ + *ε*_*a*_, where *ε*_*a*_ ∼ *N* (0, 1), that is, 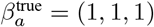. The true outcome model, i.e. the generation of *X*_1_ follows *X*_1_ = −0.5*X*_2_ + *ε*_*o*_ for continuous *X*_1_ and *X*_1_ = I(− 0.5*X*_2_ + *ε*_*o*_ *>* 0) for binary *X*_1_. In settings a) and b), the missing indicator is generated by *R* = I(*β*_*s*0_ + 0.5*Y* + 0.5*X*_2_ + 0.5*X*_3_ + *ε*_*s*_ *>* 0), where and the error terms *ε*_*s*_ and *ε*_*o*_ are correlated with *ρ* = 0 in setting a) MAR and *ρ* = 0.5 in setting b) Heckman-type MNAR. In setting c), on the other hand, the missing indicator follows a Bernoulli distribution with probability Pr(*R* = 1) = 1*/*[1 + exp( −(*β*_*s*0_ + 0.5*Y* + 0.5*X*_2_ + 0.5*X*_3_ + 0.5*X*_1_))], where the latent value of *X*_1_ itself is directly associated with its missingness, *ε*_*o*_ ∼ *N* (0, 1) and there is no *ε*_*s*_. In all settings, the selection model intercept *β*_*s*0_ is varied slightly to achieve approximately 40% missingness in *X*_1_. In terms of sites and sample size, under all three settings, we allocate a total of *N* = 1500 observations evenly (E) and unevenly (U) across *K* = 3 and 6 sites, we summarize the detailed distribution in Table 1. R code for generating the simulation results can be found at https://github.com/ly129/HDMI.

**Table 1.** Different distributions of samples in the simulation studies. Type: aggregated (A) for baseline methods, unevenly (U) and evenly (E) distributed; *K*: number of sites; *N* : total sample size; *n*^(*k*)^: sample size at site *k*.

| Type | $K$ | $N$ | $n^{(1)}$ | $n^{(2)}$ | $n^{(3)}$ | $n^{(4)}$ | $n^{(5)}$ | $n^{(6)}$ |
| --- | --- | --- | --- | --- | --- | --- | --- | --- |
| A | 1 | 1500 | 1500 |  |  |  |  |  |
| U | 3 | 1500 | 1300 | 100 | 100 |  |  |  |
| U | 6 | 1500 | 1000 | 100 | 100 | 100 | 100 | 100 |
| E | 3 | 1500 | 500 | 500 | 500 |  |  |  |
| E | 6 | 1500 | 250 | 250 | 250 | 250 | 250 | 250 |

We compare the HDMI algorithm with a number of competing algorithms, including a <u>H</u>eckman-type <u>i</u>ndependent <u>m</u>ultiple <u>i</u>mputation algorithm (HIMI) where imputation is performed locally at each site using Algorithm 1, as well as an <u>i</u>ndependent <u>m</u>ultiple <u>i</u>mputation algorithm (IMI) and two <u>d</u>istributed <u>m</u>ultiple <u>i</u>mputation algorithms (DMI using the AVGM^3^ algorithm and DMI* using the communication-efficient surrogate likelihood algorithm^5^) for MAR missingness^11^. We also provide four baselines where data aggregated across sites are imputed (if applicable) and analyzed, including the hypothetical complete data analysis (CD), complete cases analysis (CC), as well as analyses of imputed data by MI for MAR missingness (MI) and Heckman-type MI for MNAR data (HMI). In all imputation algorithm, *M* = 100 imputations are performed and the same procedure is replicated on 1000 Monte Carlo simulated datasets. Performance is evaluated by comparing the final analysis model estimates 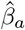 to 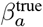, including 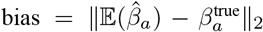, standard deviation 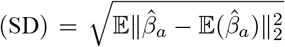 and root mean squared error 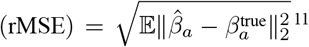. We also record the number of one-way communications (Comm) between the sites and central servers when applicable. Results are summarized in Tables 2, 3 and 4 for settings a), b) and c), respectively. We compare different methods within the same sample distribution type and number of sites (shaded blocks), particularly we compare the distributed MI algorithms with their independent counterpart, e.g. HDMI vs. HIMI, DMI vs. IMI. We also compare the results from the same methods in different blocks to determine whether they are robust against different distributions of the samples. Finally we can compare our results to those generated from imputing and analyzing hypothetical aggregated data, MI and HMI, which represent the best performance DMI and HDMI can theoretically achieve, respectively.

**Table 2.** Simulation results for setting a) one covariate is missing at random. Type: A – data is aggregated for baseline methods; U – data is unevenly distributed in *K* sites; E – data is evenly distributed in *K* sites. Method: CD – hypothetical complete data analysis; CC – complete case analysis; H – Heckman; D – distributed; I – independent. E.g. IMI: independent MI; HDMI: Heckman distributed MI.

| Type | $K$ | Method | Continuous covariate missing | | | | Binary covariate missing | | | |
| --- | --- | --- | --- | --- | --- | --- | --- | --- | --- | --- |
|  |  |  | Bias | SD | rMSE | Comm | Bias | SD | rMSE | Comm |
| A | 1 | CD | 0.000 | 0.065 | 0.065 | 0 | 0.000 | 0.084 | 0.084 | 0 |
|  | 1 | CC | 0.366 | 0.085 | 0.376 | 0 | 0.397 | 0.104 | 0.410 | 0 |
|  | 1 | MI | 0.002 | 0.078 | 0.079 | 0 | 0.003 | 0.095 | 0.095 | 0 |
|  | 1 | HMI | 0.034 | 0.132 | 0.137 | 0 | 0.040 | 0.132 | 0.138 | 0 |
| U | 3 | IMI | 0.007 | 0.078 | 0.078 | 0 | 0.005 | 0.095 | 0.095 | 0 |
|  | 3 | DMI | **0.002** | 0.078 | 0.079 | 2 | 0.006 | 0.095 | 0.095 | 2 |
|  | 3 | DMI* | **0.002** | 0.079 | 0.079 | 3 | **0.003** | 0.096 | 0.096 | 3 |
|  | 3 | HIMI | 0.090 | 0.131 | 0.159 | 0 | 0.067 | 0.129 | 0.146 | 0 |
|  | 3 | HDMI | 0.041 | 0.136 | 0.142 | 2 | 0.038 | 0.130 | 0.136 | 2 |
| U | 6 | IMI | 0.014 | 0.079 | 0.080 | 0 | 0.013 | 0.096 | 0.096 | 0 |
|  | 6 | DMI | 0.004 | 0.079 | 0.079 | 2 | 0.013 | 0.096 | 0.097 | 2 |
|  | 6 | DMI* | **0.002** | 0.079 | 0.079 | 3 | **0.003** | 0.095 | 0.095 | 3 |
|  | 6 | HIMI | 0.163 | 0.123 | 0.205 | 0 | 0.108 | 0.123 | 0.164 | 0 |
|  | 6 | HDMI | 0.050 | 0.136 | 0.144 | 2 | 0.047 | 0.123 | 0.131 | 2 |
| E | 3 | IMI | 0.006 | 0.079 | 0.079 | 0 | 0.006 | 0.095 | 0.095 | 0 |
|  | 3 | DMI | **0.002** | 0.078 | 0.078 | 2 | 0.005 | 0.095 | 0.096 | 2 |
|  | 3 | DMI* | **0.002** | 0.080 | 0.080 | 3 | **0.003** | 0.095 | 0.095 | 3 |
|  | 3 | HIMI | 0.116 | 0.130 | 0.175 | 0 | 0.117 | 0.126 | 0.172 | 0 |
|  | 3 | HDMI | 0.043 | 0.131 | 0.138 | 2 | 0.050 | 0.127 | 0.137 | 2 |
| E | 6 | IMI | 0.014 | 0.079 | 0.080 | 0 | 0.014 | 0.095 | 0.097 | 0 |
|  | 6 | DMI | 0.004 | 0.079 | 0.079 | 2 | 0.012 | 0.096 | 0.096 | 2 |
|  | 6 | DMI* | **0.002** | 0.083 | 0.083 | 3 | **0.002** | 0.096 | 0.096 | 3 |
|  | 6 | HIMI | 0.191 | 0.122 | 0.227 | 0 | 0.164 | 0.117 | 0.201 | 0 |
|  | 6 | HDMI | 0.046 | 0.127 | 0.135 | 2 | 0.061 | 0.120 | 0.135 | 2 |

**Table 3.** Simulation results for setting b) one covariate is Heckman-type missing not at random. Type: A – data is aggregated for baseline methods; U – data is unevenly distributed in *K* sites; E – data is evenly distributed in *K* sites. Method: CD – hypothetical complete data analysis; CC – complete case analysis; H – Heckman; D – distributed; I – independent. E.g. IMI: independent MI; HDMI: Heckman distributed MI.

| Type | $K$ | Method | Continuous covariate | | | | Binary covariate | | | |
| --- | --- | --- | --- | --- | --- | --- | --- | --- | --- | --- |
|  |  |  | Bias | SD | rMSE | Comm | Bias | SD | rMSE | Comm |
| A | 1 | CD | 0.000 | 0.065 | 0.065 | 0 | 0.000 | 0.084 | 0.084 | 0 |
|  | 1 | CC | 0.449 | 0.091 | 0.458 | 0 | 0.570 | 0.111 | 0.580 | 0 |
|  | 1 | MI | 0.198 | 0.086 | 0.216 | 0 | 0.147 | 0.109 | 0.183 | 0 |
|  | 1 | HMI | 0.024 | 0.115 | 0.117 | 0 | 0.022 | 0.121 | 0.123 | 0 |
| U | 3 | IMI | 0.195 | 0.086 | 0.213 | 0 | 0.150 | 0.108 | 0.185 | 0 |
|  | 3 | DMI | 0.199 | 0.086 | 0.217 | 2 | 0.144 | 0.108 | 0.181 | 2 |
|  | 3 | DMI* | 0.198 | 0.086 | 0.216 | 3 | 0.148 | 0.109 | 0.184 | 3 |
|  | 3 | HIMI | 0.068 | 0.114 | 0.133 | 0 | 0.052 | 0.121 | 0.131 | 0 |
|  | 3 | HDMI | **0.030** | 0.120 | 0.123 | 2 | **0.028** | 0.125 | 0.128 | 2 |
| U | 6 | IMI | 0.191 | 0.086 | 0.209 | 0 | 0.155 | 0.107 | 0.189 | 0 |
|  | 6 | DMI | 0.200 | 0.086 | 0.218 | 2 | 0.141 | 0.110 | 0.178 | 2 |
|  | 6 | DMI* | 0.198 | 0.086 | 0.216 | 3 | 0.147 | 0.108 | 0.183 | 3 |
|  | 6 | HIMI | 0.130 | 0.113 | 0.172 | 0 | 0.096 | 0.121 | 0.155 | 0 |
|  | 6 | HDMI | **0.038** | 0.125 | 0.131 | 2 | **0.046** | 0.125 | 0.133 | 2 |
| E | 3 | IMI | 0.195 | 0.086 | 0.213 | 0 | 0.151 | 0.108 | 0.185 | 0 |
|  | 3 | DMI | 0.199 | 0.086 | 0.217 | 2 | 0.145 | 0.108 | 0.181 | 2 |
|  | 3 | DMI* | 0.198 | 0.089 | 0.217 | 3 | 0.148 | 0.108 | 0.183 | 3 |
|  | 3 | HIMI | 0.078 | 0.115 | 0.139 | 0 | 0.092 | 0.127 | 0.157 | 0 |
|  | 3 | HDMI | **0.032** | 0.119 | 0.123 | 2 | **0.039** | 0.123 | 0.129 | 2 |
| E | 6 | IMI | 0.191 | 0.086 | 0.209 | 0 | 0.155 | 0.107 | 0.188 | 0 |
|  | 6 | DMI | 0.200 | 0.086 | 0.218 | 2 | 0.141 | 0.109 | 0.178 | 2 |
|  | 6 | DMI* | 0.198 | 0.093 | 0.218 | 3 | 0.148 | 0.109 | 0.184 | 3 |
|  | 6 | HIMI | 0.146 | 0.116 | 0.186 | 0 | 0.143 | 0.121 | 0.187 | 0 |
|  | 6 | HDMI | **0.042** | 0.121 | 0.129 | 2 | **0.062** | 0.120 | 0.135 | 2 |

**Table 4.** Simulation results for setting c) one covariate is non-Heckman missing not at random. Type: A – data is aggregated for baseline methods; U – data is unevenly distributed in *K* sites; E – data is evenly distributed in *K* sites. Method: CD – hypothetical complete data analysis; CC – complete case analysis; H – Heckman; D – distributed; I – independent. E.g. IMI: independent MI; HDMI: Heckman distributed MI.

| Type | $K$ | Method | Continuous covariate | | | | Binary covariate | | | |
| --- | --- | --- | --- | --- | --- | --- | --- | --- | --- | --- |
|  |  |  | Bias | SD | rMSE | Comm | Bias | SD | rMSE | Comm |
| A | 1 | CD | 0.003 | 0.066 | 0.066 | 0 | 0.003 | 0.083 | 0.083 | 0 |
|  | 1 | CC | 0.212 | 0.090 | 0.230 | 0 | 0.381 | 0.117 | 0.399 | 0 |
|  | 1 | MI | 0.315 | 0.086 | 0.326 | 0 | 0.150 | 0.112 | 0.187 | 0 |
|  | 1 | HMI | 0.049 | 0.113 | 0.123 | 0 | 0.088 | 0.172 | 0.193 | 0 |
| U | 3 | IMI | 0.311 | 0.087 | 0.323 | 0 | 0.152 | 0.111 | 0.188 | 0 |
|  | 3 | DMI | 0.316 | 0.087 | 0.327 | 2 | 0.148 | 0.112 | 0.185 | 2 |
|  | 3 | DMI* | 0.315 | 0.087 | 0.327 | 3 | 0.150 | 0.111 | 0.187 | 3 |
|  | 3 | HIMI | 0.096 | 0.117 | 0.151 | 0 | 0.122 | 0.160 | 0.201 | 0 |
|  | 3 | HDMI | **0.055** | 0.120 | 0.132 | 2 | **0.078** | 0.181 | 0.197 | 2 |
| U | 6 | IMI | 0.307 | 0.087 | 0.319 | 0 | 0.155 | 0.111 | 0.191 | 0 |
|  | 6 | DMI | 0.317 | 0.087 | 0.329 | 2 | 0.145 | 0.112 | 0.184 | 2 |
|  | 6 | DMI* | 0.315 | 0.087 | 0.327 | 3 | 0.150 | 0.111 | 0.187 | 3 |
|  | 6 | HIMI | 0.161 | 0.118 | 0.200 | 0 | 0.171 | 0.147 | 0.225 | 0 |
|  | 6 | HDMI | **0.068** | 0.131 | 0.147 | 2 | **0.091** | 0.174 | 0.196 | 2 |
| E | 3 | IMI | 0.312 | 0.087 | 0.323 | 0 | 0.152 | 0.111 | 0.188 | 0 |
|  | 3 | DMI | 0.316 | 0.086 | 0.327 | 2 | 0.148 | 0.111 | 0.185 | 2 |
|  | 3 | DMI* | 0.315 | 0.088 | 0.327 | 3 | 0.150 | 0.111 | 0.186 | 3 |
|  | 3 | HIMI | 0.108 | 0.123 | 0.163 | 0 | 0.182 | 0.148 | 0.235 | 0 |
|  | 3 | HDMI | **0.062** | 0.126 | 0.141 | 2 | **0.115** | 0.152 | 0.191 | 2 |
| E | 6 | IMI | 0.307 | 0.087 | 0.319 | 0 | 0.155 | 0.111 | 0.191 | 0 |
|  | 6 | DMI | 0.317 | 0.087 | 0.329 | 2 | 0.145 | 0.112 | 0.183 | 2 |
|  | 6 | DMI* | 0.316 | 0.092 | 0.329 | 3 | 0.151 | 0.112 | 0.188 | 3 |
|  | 6 | HIMI | 0.181 | 0.122 | 0.218 | 0 | 0.224 | 0.132 | 0.260 | 0 |
|  | 6 | HDMI | **0.082** | 0.127 | 0.151 | 2 | **0.133** | 0.156 | 0.205 | 2 |

The results for the MAR case in setting a) are summarized in Table 2. The Heckman MI algorithms are outperformed by their non-Heckman counterparts across the board, i.e. lower bias as well as SD and rMSE. These may be viewed as a small price that the more complex Heckman MI algorithms have to pay to be able to impute MNAR data. Nonetheless, HDMI is able to provide significant improvements over HIMI and are much more comparable to the baseline (HMI in the table). Similarly, DMI and DMI* are able to generate results that are close to those acquired by (hypothetically) imputing and analyzing the aggregated data (MI in the table), while marginally outperform the non-distributed IMI. These suggest that, unlike IMI/HIMI, the distributed DMI/HDMI are not sensitive to the number of sites or how the observations are distributed across sites, suggesting the distributed imputation’s robustness against small sample sizes at some sites and uneven distributions across sites. Overall, under the MAR setting, although outperformed by MI methods developed for MAR, our HDMI algorithm can provide decent results (around 5% bias), and more importantly, the much needed improvements over independent local Heckman imputation using existing methods^12^.

Under the Heckman-type MNAR setting b)^12^, multiple imputation methods designed for MAR missingness no longer suffice and Heckman MI algorithms become necessary (Table 3). The best performer HDMI can achieve under 5% bias in most cases, which is comparable to the hypothetical non-distributed HMI. In comparison, the HIMI is generating much higher bias (5% to 15%) than HDMI, justifying the need for distributed imputation for MNAR data. Finally, similar to Table 2, HDMI is less sensitive to how the data is distributed than HIMI, including the number of sites and the sample sizes of each site. Last but not least, we observe that the biases are similar in the Heckman-type MNAR setting (*ρ* = 0.5) to (if not better than) those in the MAR setting (*ρ* = 0) in Table 2, suggesting that the Heckman-based HDMI and HIMI are robust against the correlation coefficient *ρ*, which quantifies how far the missingness deviates from MAR, or how not at random the missingness is. On the other hand, for Non-Heckman MI, IMI, DMI and DMI*, the bias in some cases can be as high as 20%, which is substantial comparing to those under 1% under the MAR setting in Table 2. These results suggest that non-Heckman MI algorithms are very sensitive to Heckman-type MNAR and can lead to significant bias.

Using setting c), we test the robustness of the HDMI algorithm against Non-Heckman MNAR and summarize the results in Table 4. The non-Heckman MNAR, where the latent value of the missing covariate itself directly affects its missingness (likely a more intuitive mechanism than the Heckman MNAR), poses a greater challenge for the HDMI algorithm, as well as the competing MI algorithms. When the missing covariate is a continuous variable, the HDMI is able to guarantee *<*10% bias, while the value can be as high as 13% when the missing covariate is binary. In contrast, the non-Heckman imputation methods (MI, IMI, DMI, DMI*) yield roughly 30% and 15% bias for the continuous and binary missing variable, respectively. In addition, the non-distributed HIMI also generates much higher bias than the HDMI. That said, the HDMI clearly outperforms all other methods across the board, in both even and uneven distribution types and different number of sites. Finally, with only two one-way communications between the central server and sites, the HDMI can generate imputation results that are comparable to the centralized HMI. These make the HDMI the best option at our hands to impute missing data under distributed settings that are believed to be MNAR.

### Real-world data analysis

We perform a real-world case study using the GCAS. In addition to the introduction in the previous sections, the GCASR is a program implemented to reduce morbidity, mortality and disability due to stroke, the incidence of recurrent stroke, and stroke-related disparity in Georgia (https://dph.georgia.gov/stroke/georgia-coverdell-acute-stroke-registry). The program encourages collaboration among EMS providers, hospitals and other institutions in Georgia to improve stroke care quality. For demonstration purpose, we investigate the association between arrival-to-CT time and EMSNote, adjusting for potential confounders Weekend. To perform this analysis, we include patients arrived at the hospital from home or scene by EMS only. In addition, due to the highly right-skewed distribution, we select only patients with arrival-to-CT time under ten hours and perform a (natural) log transformation. By doing these, we are able to keep over 96.2% of total patients and exclude potential outliers with high values based on inspections of the distribution of arrival-to-CT time, and transform it from a skewed variable to one that is more approximately normal. The binary EMSNote variable is subject to missingness and the proportion of missingness is highly discrepant across hospitals (Figure 1). We excluded hospitals that have under 5% of missingness in EMSNote or less than 50 observations. This exclusion is due to an important limitation of the Heckman models, that is its numerical instability when the missing proportion is close to either zero or one and when the sample size of some sites are small. We will elaborate on it in the Discussion and provide a workaround. Finally, as per the exclusion-restriction rule of the Heckman model, we include a binary covariate EducEMS in the selection model to predict the missingness of EMSNote. This covariate indicates whether there have been documentation that the patient and/or caregiver received education and/or resource materials regarding how to activate EMS for stroke, which has been shown to be an important factor in stroke care^22^. In the end, after excluding patients with missing information in variables other than EMSNote, we are able to assemble a dataset containing 4398 patients distributed in seven hospitals.

As the underlying true value of the regression coefficients are unknown, the goal of this analysis is to show that, through a simplified real-world case study, the choice of imputation method can have significant impact on the estimation results. Using the same method acronyms, we perform CC, MI and HMI on the aggregated data, as well as IMI, DMI, DMI*, HIMI and HDMI on the data as if it was distributed. We summarize the estimates in Table 5. In the CC analysis, EMSNote is associated with 0.24 increase in the log of arrival-to-CT time, after adjusting for potential confounders including Gender and Weekend. If we perform HDMI to impute the missing values in EMSNote, the association becomes 0.16. Comparing the imputation algorithms, the methods for MNAR based on the Heckman model generate more consistent results than those for the imputation of MAR data. In addition, the MI, IMI and DMI, comparing to their Heckman counterparts HMI, HIMI and HDMI, lead to estimates that are closer to those of the complete case analysis. A possible explanations for the closeness between the MI, IMI, DMI and CC is that EMSNote is MCAR in which case all the estimates are relatively unbiased. However, in case there is strong evidence or reasoning against the MCAR assumption, then the majority of MI methods suggest that the latent missing mechanism is causing an overestimation, by the complete case analysis, of the association between EMSNote and the arrival-to-CT time. More comprehensive studies are needed to verify the association between EMSNote and arrival-to-CT time. We discuss important practical considerations associated with the application of the HDMI algorithm in the section below.

**Table 5.** Association between the logarithm arrival-to-CT time and advance notification by EMS, adjusting for gender and day of arrival estimated by different multiple imputation algorithms.

|  | Intercept | EMSNote | Gender | Weekend |
| --- | --- | --- | --- | --- |
| CC | 3.62 | 0.24 | 0.07 | −0.03 |
| MI | 3.56 | 0.28 | 0.06 | −0.05 |
| HMI | 3.60 | 0.13 | 0.06 | −0.04 |
| IMI | 3.59 | 0.23 | 0.06 | −0.04 |
| DMI | 3.61 | 0.18 | 0.06 | −0.04 |
| DMI* | 3.41 | 0.59 | 0.06 | −0.06 |
| HIMI | 3.61 | 0.16 | 0.06 | −0.04 |
| HDMI | 3.62 | 0.15 | 0.06 | −0.04 |

## Discussion and Conclusions

Through extensive numerical experiments, we show that the HDMI algorithm proposed in this paper outperforms existing distributed MI algorithms designed for MAR data when the missing mechanism is MNAR. In addition, when the missing mechanism is MAR, the HDMI exhibits decent performance comparing to algorithms that are specifically designed to impute MAR missing data. This suggests that the HDMI is robust against the unknown missing mechanism, therefore can be particularly useful when some of the sites show evidence of MNAR. Furthermore, the Heckman model, as well as the HDMI, can provide some evidence of the missing mechanism in practice. Particularly, the model involves the estimation of the correlation coefficient *ρ* that quantifies how not at random the missingness is. For example, in multi-center clinical studies, it can be difficult to decide whether losses to follow-up in some patients are MAR or MNAR^23^. One could argue that patients who experience worsening conditions are likely to drop out (e.g. worse psychological conditions), or on the contrary, patients whose conditions have improved may not feel the need to continue to seek care. The value of the estimated correlation coefficient can give practitioners some sense of the missing mechanism. In addition, the HDMI features efficient communication and preservation of data privacy, making it the ideal method for imputation of missing data in distributed EHRs. Our proposed algorithm is subject to an important limitation. In our experiments, particularly the real-world example, we experience convergence issues when the missing rate is close to either 0 or 1, or when the sample size at some sites are very small. However, there is a solution – for such sites that cannot provide much information on the missing mechanism and/or missing value, we can exclude them from the first step of the HDMI algorithm, that is, we do not fit a Heckman model using data at these sites. This does not prevent us from using the Heckman model coefficients averaged across other sites to perform imputation at these sites. Researchers can borrow information across sites to improve the imputation results for sites with very high missing rate or limited sample sizes, potentially improving the fairness for certain minority groups. In fact, this is one of the motivations for distributed MI discussed in the introduction. Now, we list a few important practical considerations when HDMI is applied to distributed EHRs. First, we may want to decide for which variables HDMI should be applied due to probable MNAR, and for which variables simpler MAR imputation algorithms will suffice. Then for each variable that is believed to be MNAR, we identify sites with moderate missing proportion and healthy sample size through a priori planning and coordination with the sites. Next, for each variable imputed with HDMI, we want to carefully select at least one supplementary variable, as per the restriction-exclusion rule based on domain-specific knowledge. In summary, the HDMI is designed for and is capable of performing effective imputation of MNAR data, and is able to adapt to MAR data for distributed EHRs and other data. The HDMI provides a robust and reliable addition to existing distributed MI algorithms if there is evidence of MNAR. To our knowledge, the HDMI is the only distributed imputation algorithm for MNAR data in the literature and it can be readily used in real-world applications.

## Data Availability

The real data analyzed in this article were provided by the Georgia Coverdell Acute Stroke Registry and restrictions apply to the availability of these data. Request for access to the data should be submitted to and approved by the Georgia Coverdell Acute Stroke Registry.

## Acknowledgements

This work was supported by the National Institutes of Health, U01-CA274576. The content is solely the responsibility of the authors and does not necessarily represent the official views of the National Institutes of Health. XJ is CPRIT Scholar in Cancer Research (RR180012), and he was supported in part by Christopher Sarofim Family Professorship, UT Stars award, UTHealth startup, the National Institute of Health (NIH) under award number R01AG066749, R01AG066749-03S1, R01LM013712, R01LM014520, R01AG082721, R01AG066749, U01AG079847, U24LM013755, U01CA274576, U54HG012510, 1OT2OD032581-02-420, 1OT2OD032581-02-211, 1OT2OD032581-02-164, OT2OD032701 and the National Science Foundation (NSF) #2124789.

